# Heterogeneity of Provider Preferences for HIV Care Coordination Program Features: Latent Class Analysis of a Discrete Choice Experiment

**DOI:** 10.1101/2022.04.26.22274351

**Authors:** Chunki Fong, Madellena Conte, Rebecca Zimba, Jennifer Carmona, Gina Gambone, Abigail Baim-Lance, McKaylee Robertson, Mary Irvine, Denis Nash

## Abstract

**Background:** The PROMISE study was launched in 2018 to assess revisions to an HIV care coordination program (CCP) designed to address gaps in care and treatment engagement among people living with HIV in New York City (NYC). We report on the heterogeneity of provider preferences regarding a revised CCP elicited from a discrete choice experiment (DCE).

**Methods:** From January to March 2020, 152 CCP providers in NYC completed a DCE with 4 program attributes: 1) help with adherence to antiretroviral therapy, 2) help with primary care appointments, 3) help with issues other than primary care, and 4) program visit location. Each attribute had 3-4 levels. Latent class analysis (LCA) was used to detect subgroups with differing attribute importance and part-worth utility patterns. Choice simulation was used to estimate providers’ endorsement of eight hypothetical CCPs.

**Results:** LCA identified three subgroups. The two larger subgroups (n = 133) endorsed more intensive attribute levels, particularly clients receiving directly observed therapy, and home visits. The remaining smaller subgroup (n = 19) endorsed clients receiving medication reminders and meeting with clients at the program. Simulation showed that intensive medical case management programs had the highest degree of endorsement (62%).

**Conclusion:** While our results indicate high endorsement among providers for intensive CCP features, overall, they also suggest the need for flexible service delivery options to meet the needs of the clients that these programs serve. Additional information sharing across and within agencies may be warranted to improve the fidelity with which the CCP is implemented.

## Introduction

Disparities in HIV viral suppression (VS) among people with HIV (PWH) in New York City (NYC) constitute a major obstacle to achieving the goals of the Ending the HIV Epidemic initiative [1]. In 2019, 77% of PWH in NYC had VS, though inequities persisted with lower rates of VS among Black and Latino people, transgender individuals, young people, and people with a history of injection drug use [2,3]. To address these disparities and strengthen the HIV care continuum as a whole, the NYC Department of Health and Mental Hygiene (NYC Health Department) launched a multi-component HIV Care Coordination Program (CCP) in 2009 directed toward the most vulnerable, high-need PWH in NYC [4]. The CCP combined several evidence-based elements including antiretroviral therapy (ART) adherence support, case management, outreach for initial case finding, and patient navigation [5]. Despite the success of this CCP as measured by improved VS for CCP participants newly diagnosed or consistently unsuppressed, the absence of an effect on VS among previously diagnosed PWH with any suppression in the year prior to enrollment left ample room for improvement [5,6]. Based on lessons learned in the first several years of the program’s existence, the NYC Health Department undertook a substantial revision of the CCP model and a resolicitation of CCP service delivery contracts in 2017-2018, which led to its designation by Centers for Disease Control (CDC) as a Structural Evidence-Based Intervention [7]. The salient revisions to the CCP included removing rigid program tracks, the optional use of video chat as a mode of delivery for services such as directly observed therapy (DOT), counseling for client HIV self-management capacity, and fee-for-service reimbursement model that accounts for staff travel to and from clients’ homes [8]. The PROMISE study (<u>P</u>rogram <u>R</u>efinements to <u>O</u>ptimize <u>M</u>odel Impact and <u>S</u>calability based on <u>E</u>vidence) was launched in 2018 to assess the impact and the implementation of the revised program model, as compared to the original CCP [8].

Discrete Choice Experiments (DCE) are research tools that quantify preferences and trade-offs in decision making and are increasingly applied to HIV care program design [9]. Provider and client preferences of CCP features can guide future revisions to program design and ultimately optimize real-world effectiveness. While past research has explored client preferences for HIV care and treatment services [10–12], little work has been done to systematically ascertain the preferences of providers. Attention to provider preferences, including potential heterogeneity of those preferences, could improve the acceptability, feasibility, sustainability, and implementation fidelity of care and treatment programs in the settings where they will be delivered [13,14]. In this way, a better understanding of provider perspectives on program implementation and delivery approaches has the potential to result in improvements in patient outcomes. We report on the results of a DCE among NYC CCP providers to understand the preference heterogeneity in this population with regard to specific attributes of the revised CCP.

## Methods

### Study setting and population

Ryan White Part A funding in NYC funds services other than medical care. Therefore, our target population included non-medical providers in the core CCP positions of patient navigators/health educators, care coordinators/case managers, and program directors or other administrators at any of the 25 agencies implementing the revised CCP. All staff in those core program roles were eligible to participate. Of the 25 agencies, 10 agencies were community health centers, 6 were private hospitals, 3 were public hospitals, and 6 were community-based organizations. Eleven agencies were located in Brooklyn, 10 were located in Manhattan, 9 were located in the Bronx, 4 were located in Queens, and 1 was located in Staten Island. The study was approved by the NYC Health Department Institutional Review Board, which is the IRB of record for the PROMISE study. All participants provided informed consent electronically to participate in the study.

### DCE Design

Lighthouse Studio Version 9.8.1. (Sawtooth Software, Provo, Utah, USA) was used to construct and implement the DCE, specifically the Choice-Based Conjoint (CBC) question format. Here we briefly summarize the details of the DCE design, which have been previously described [15]. The DCE included four attributes as listed in Table 1: two attributes described with four levels and two described with three levels. All attribute levels were accompanied with images to improve understanding and ease the task of comparing hypothetical care coordination designs. Attributes and levels were developed from client and provider focus studies, and with feedback from the study’s advisory board and subject area experts on the study team. Preferences for care coordination features were solicited by presenting respondents with 10 choice tasks, a number that both minimizes cognitive burden and maximizes the efficiency of the DCE design [16]. Each choice task presented respondents with two models of the CCP that differed by one or more of the study’s attribute levels, requiring respondents to make a series of trade-offs between the attribute levels. In each choice task, the respondent was asked for their preference

**Table 1:**
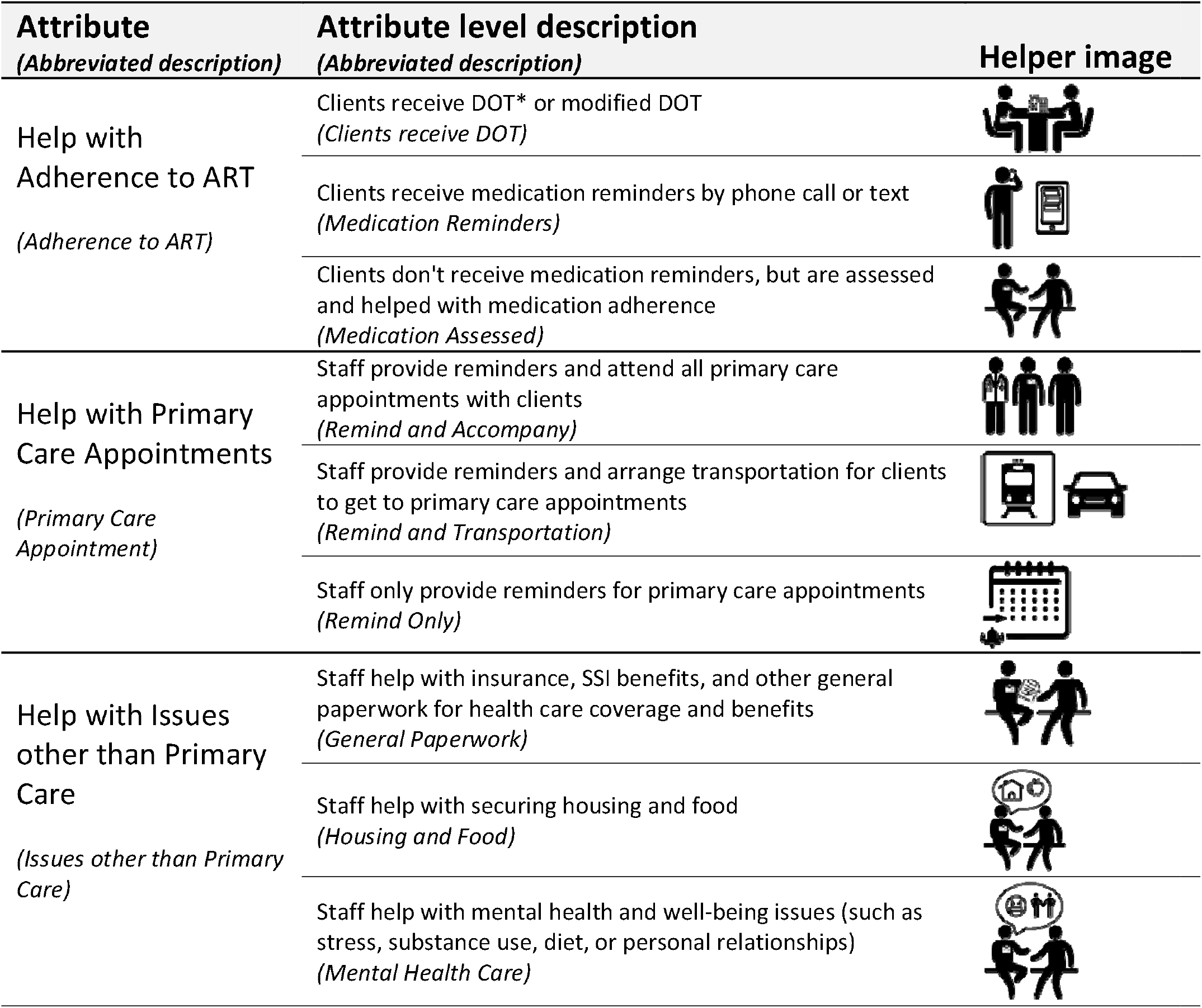
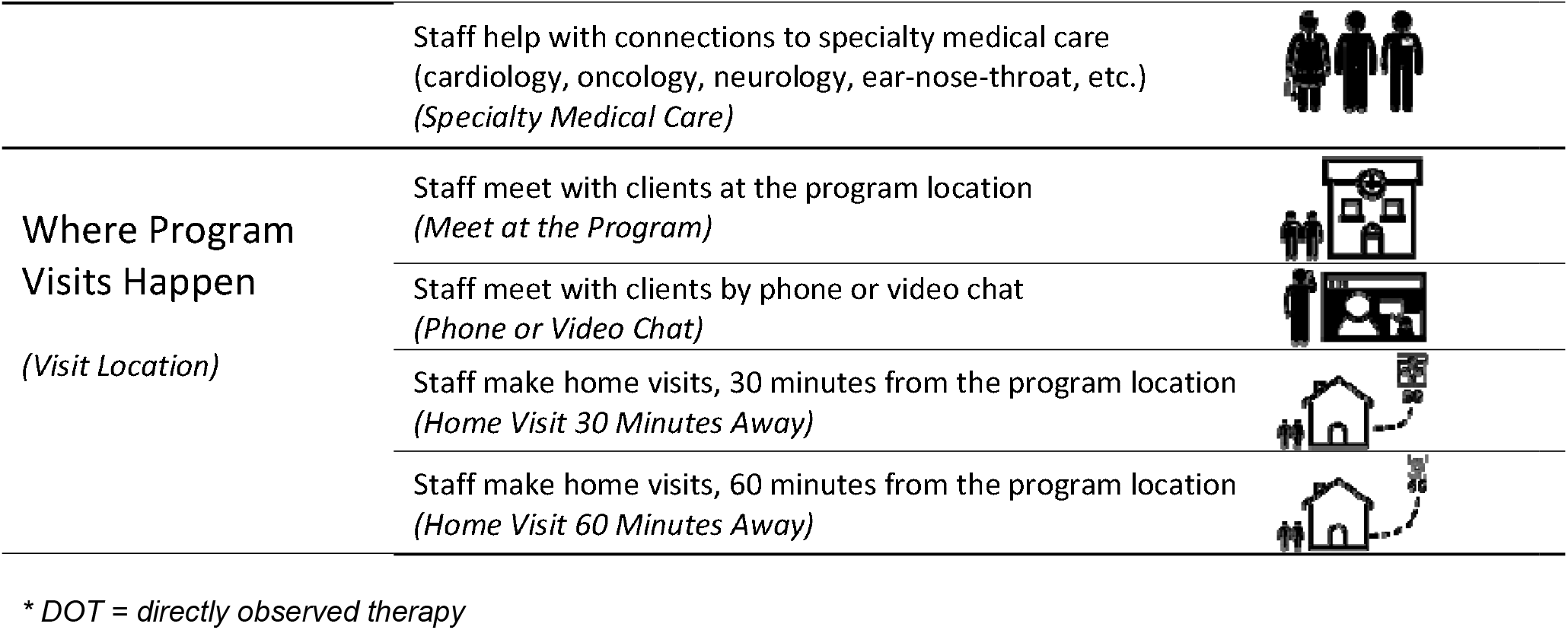
Attributes and levels in the DCE exploring preferences for HIV care coordination services in New York City.

| Attribute<br>(Abbreviated description) | Attribute level description<br>(Abbreviated description) | Helper image |
| --- | --- | --- |
| Help with Adherence to ART<br>(Adherence to ART) | Clients receive DOT* or modified DOT<br>(Clients receive DOT) |  |
|  | Clients receive medication reminders by phone call or text<br>(Medication Reminders) |  |
|  | Clients don't receive medication reminders, but are assessed and helped with medication adherence<br>(Medication Assessed) |  |
| Help with Primary Care Appointments<br>(Primary Care Appointment) | Staff provide reminders and attend all primary care appointments with clients<br>(Remind and Accompany) |  |
|  | Staff provide reminders and arrange transportation for clients to get to primary care appointments<br>(Remind and Transportation) |  |
|  | Staff only provide reminders for primary care appointments<br>(Remind Only) |  |
| Help with Issues other than Primary Care<br>(Issues other than Primary Care) | Staff help with insurance, SSI benefits, and other general paperwork for health care coverage and benefits<br>(General Paperwork) |  |
|  | Staff help with securing housing and food<br>(Housing and Food) |  |
|  | Staff help with mental health and well-being issues (such as stress, substance use, diet, or personal relationships)<br>(Mental Health Care) |  |
|  | Staff help with connections to specialty medical care (cardiology, oncology, neurology, ear-nose-throat, etc.)<br>(*Specialty Medical Care*) |  |
| Where Program Visits Happen<br>(*Visit Location*) | Staff meet with clients at the program location<br>(*Meet at the Program*) |  |
|  | Staff meet with clients by phone or video chat<br>(*Phone or Video Chat*) |  |
|  | Staff make home visits, 30 minutes from the program location<br>(*Home Visit 30 Minutes Away*) |  |
|  | Staff make home visits, 60 minutes from the program location<br>(*Home Visit 60 Minutes Away*) |  |

\* DOT = directly observed therapy

“Imagine that you had to choose between two programs with the features below. Select the one that you would prefer.” The design did not include a None option.

Sawtooth Software’s experimental design module generated 300 different versions of the survey that were balanced (across choice tasks, each attribute level appeared with the same frequency) and orthogonal (each pair of attribute levels appeared the same number of times across all pairs of attributes in the design) [17,18]. The standard error for each level ranged from 0.045 - 0.059 and the design’s relative D-efficiency was 88%, compared to a completely enumerated design.

### Sample Size

The minimum sample size to estimate main effects was estimated to be 100 respondents using the formula “n ≥ 500c/ta,” where ‘n’ is the number of respondents, ‘t’ is the number of choice tasks, ‘a’ is the number of options per choice task, and ‘c’ is largest number of levels for any attribute [19]. We set our target sample size at 150 respondents.

### Data Collection

In January 2020, we emailed individual links with survey IDs to the online survey to 227 providers from the 25 revised CCP implementing agencies. The DCE could be completed in any modern browser on a desktop or laptop computer, tablet, or phone. Following the DCE tasks, we included questions to elicit data about providers’ age, race, ethnicity, gender identity, and length of time providing care coordination services. Participants were compensated with $25 gift cards upon completion of the survey. The survey was closed in early March 2020, before the first wave of the COVID-19 pandemic in NYC. In the final dataset we merged additional staff and agency descriptive data, such as staff role, agency location, and CCP budget, gathered from program liaisons and existing NYC Health Department contract records.

### Analysis

#### Latent Class Analysis

The Latent Class Analysis (LCA) method in Lighthouse Studio’s Analysis Manager was implemented to understand the heterogeneity in providers’ preferences. LCA detects subgroups with differing preferences through patterns in estimates of part-worth utilities and of relative attribute importances. Part-worth utilities are estimates of preference weights for each attribute level based on how often those levels were present in the care coordination service packages chosen by each respondent. The higher the utility, the more often the level is chosen or preferred. The relative importance of an attribute is the proportion of the range of utilities for the attribute to the sum of ranges for all attributes. It describes the observed influence of each attribute on the respondents’ decision making [20]. The higher the percentage of relative importance, the more impactful the choice of level within the attribute was on choice. Sample size may differ among the subgroups detected by LCA, depending on the heterogeneity of the sample.

Akaike’s information criterion (AIC), Log-likelihood ratio, and the Relative Chi-Square difference were the model statistics that guided the selection of the optimal LCA solution for our data [13,21]. Based on the recommended default setting, solutions for two groups up to five groups were compared. Part-worth utilities and relative attribute importances were compared across the subgroups. We used SAS software (Release 9.4 SAS Institute Inc., Cary, NC, USA) for additional analyses.

#### Choice Simulation Analysis

In addition to estimating part-worth utilities and relative importances across the subgroups, we used Sawtooth Software’s Choice Simulator to estimate “shares of preference”: the proportion of providers within each group that would select hypothetical programs. The hypothetical programs, as listed and described in Table 2, were designed by experts on care coordination program design from the NYC Health Department and assembled using the attributes and levels from the DCE. The final set included eight distinct programs which varied in service intensity and were grouped into three broad categories: basic medical management, medium medical case management, and intensive medical case management. Our simulation scenario was designed to more accurately reflect the landscape of CCP implementation, with flexible options for supporting clients in each of the attribute areas available to providers. For example, in the DCE help with specialty medical care and help with mental health and well-being could never appear in the same model of the choice task, though in reality, these could both be offered to clients at the same time. Our choice simulation analysis allowed us to mimic such a scenario.

**Table 2:**
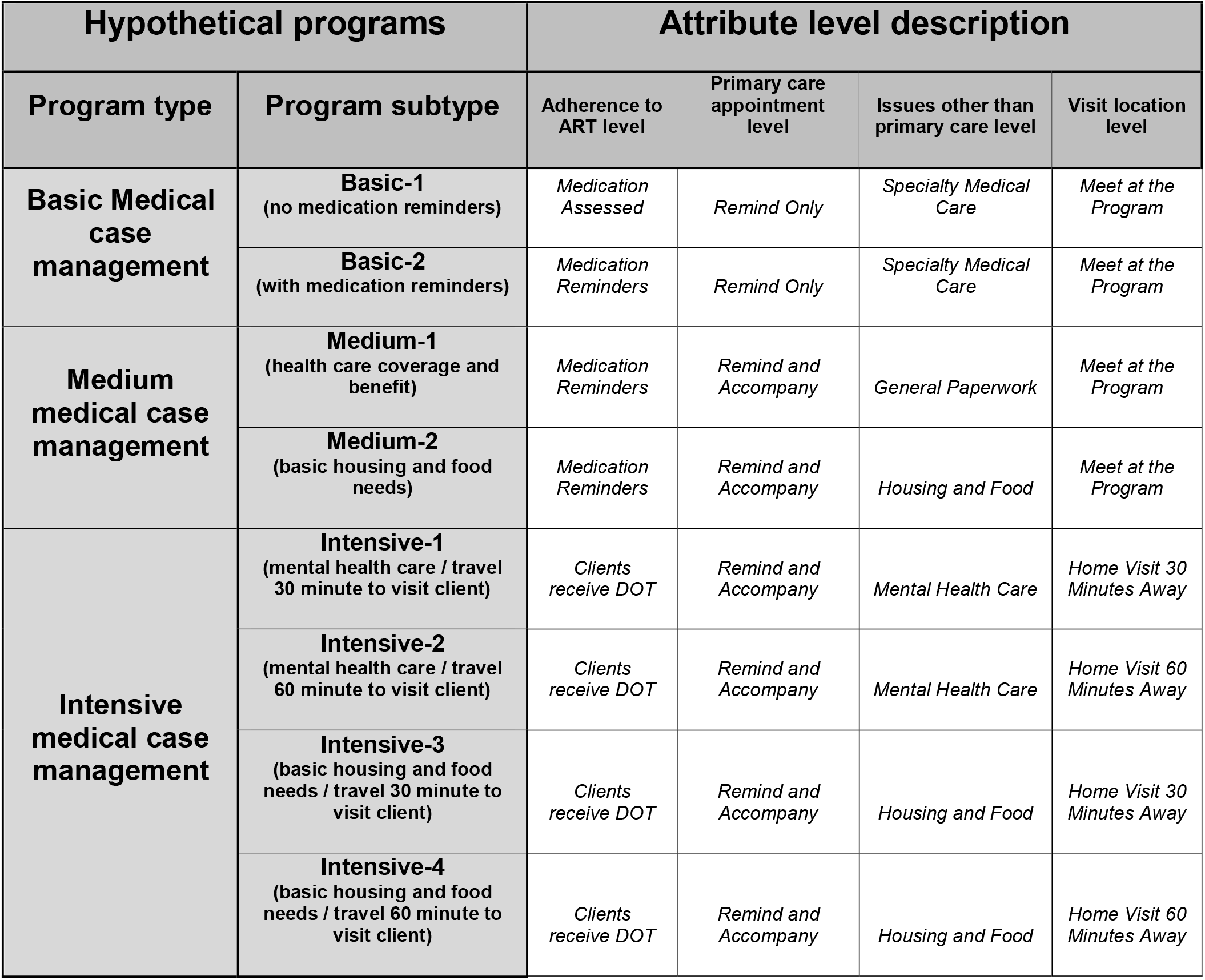
List of hypothetical programs with the corresponding attribute levels.

Using the utilities derived from the DCE surveys, the choice simulation package predicted the types of whole programs, rather than individual program features, that groups of providers would be more likely to endorse. We used the randomized first choice model, in which utilities are summed across the levels in each hypothetical option and exponentiated to determine the probability of an individual preferring one hypothetical option over the others. Randomized first choice has a stronger predictive ability compared to other simulation methods, and accounts both for random error in the point estimates of utilities and similarity within the hypothetical options [22,23].

## Results

### Characteristics of provider respondents & group segmentation

Table 3 describes the characteristics of the 152 providers from the 35 agencies implementing the CCP who participated in the study, stratified by preference group. Overall, 79 (52%) of the participating providers were 20-39 years old, 125 (82.3%) were Latino/a or Black, and 104 (68.4%) identified as female. Most of the providers (99, 65.1%) were navigator-type staff, with 33 (21.7%) care coordinator-type staff, and 20 (13.2%) administrative staff. Over half of the providers (88, 57.9%) had been providing care coordination services for more than 2 years.

**Table 3:**
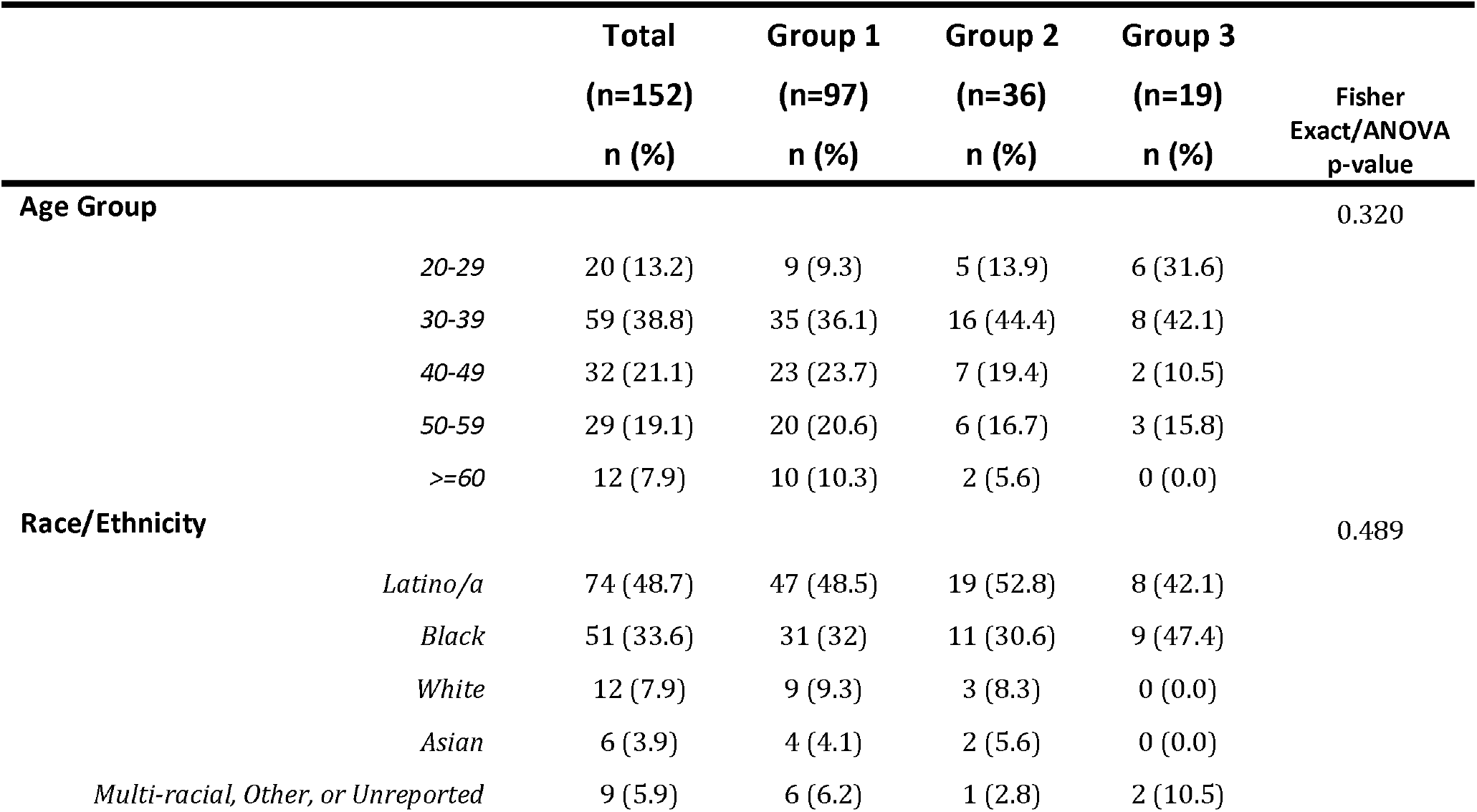
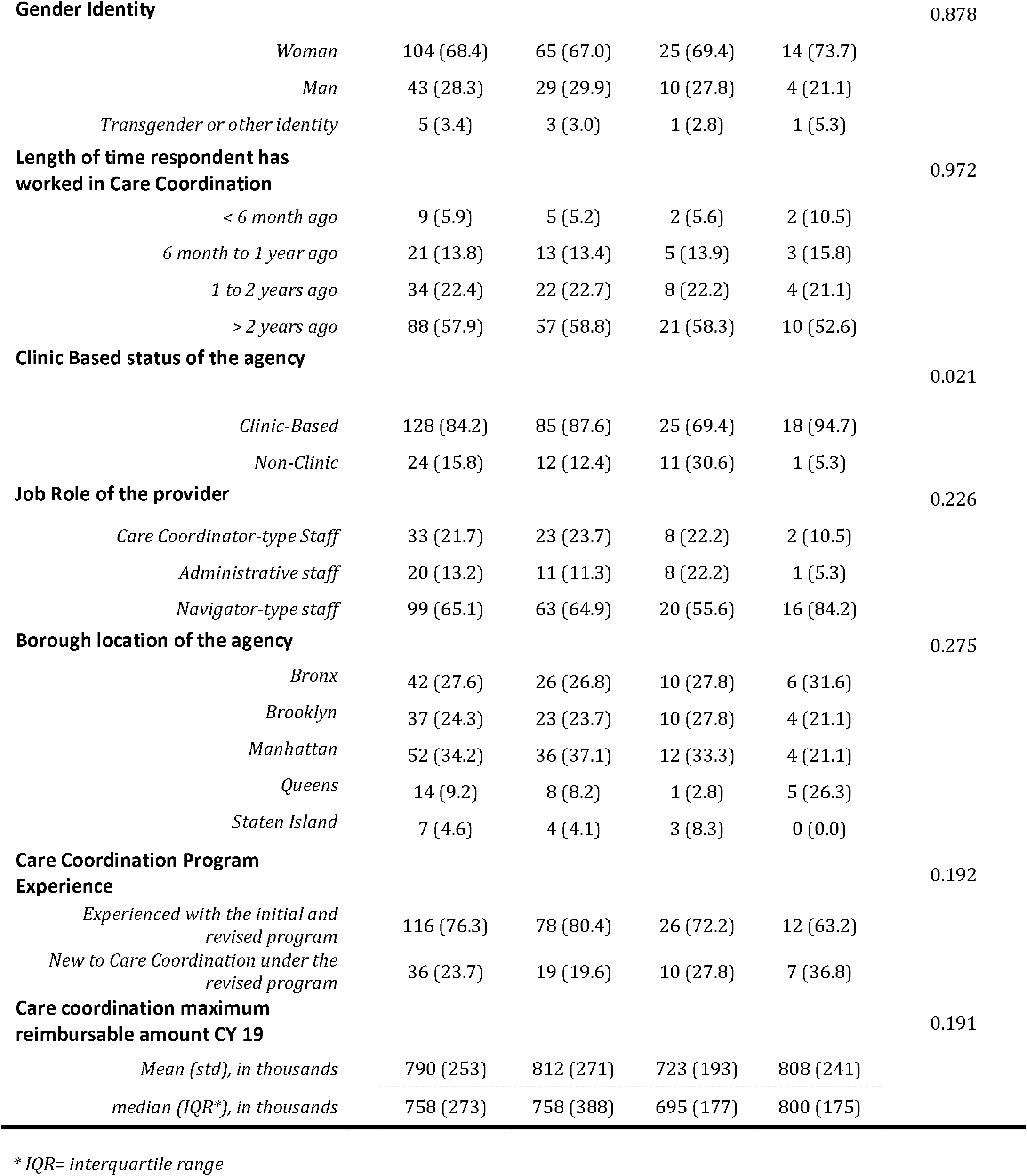
Characteristics of provider respondents.

In terms of agency characteristics, 128 (84%) providers were from agencies that are clinic-based, and 116 (76.3%) were from agencies experienced with both the initial and revised CCP. While many of the participating agencies had CCP service sites in more than one borough, 52 providers (34.2%) were from a site located in Manhattan, 42 providers (27.6%) were from a site located in the Bronx, and 37 providers (24.3%) were from a site located in Brooklyn.

To determine the appropriate number of latent class groups we examined the AIC, Log-likelihood ratio, and the Relative Chi-Square difference for the two to five group solutions. Though the 5-class solution had a lower AIC and a higher Log-likelihood, we chose to describe the model as a 3-class solution because of a higher Chi-Square difference, and for its simplicity. See Supplemental Materials for more details.

The 3-class LCA solution yielded 97 members (63.8%) in Group 1, 36 members (23.7%) in Group 2, and 19 members (12.5%) in Group 3. Agency type (Clinic-based vs non-clinic-based) was the only characteristics that yielded statistical significance among the three groups. Group 3 had a higher proportion of providers who worked at a clinic-based CCP (94.7%) compared to Group 1 (87.6%) and Group 2 (69.4%). Though not statistically significant, other differences worth noting are that Group 3 providers were more likely to work at an agency with experience only with the revised CCP (36.8%) than providers in Group 1 or Group 2 (19.6% and 27.7%, respectively). Group 3 providers were also more likely to be navigator-type staff (84.2%), as compared with providers in Group 1 and Group 2 (64.9% and 55.6%, respectively). Finally, providers in Group 1 and Group 2 were more likely to work at agencies in Manhattan (37.1% and 33.3% respectively), compared with providers in Group 3 who tended to be located at agencies in the Bronx (31.6%) or Queens (26.3%).

### Relative Importance and Part-Worth Utilities

Figure 1 compares the attribute relative importances across the three groups and Table 4 compares the part-worth utilities. Figure 2 is a graphical representation of Table 4 to display the patterns and the ranges of the part-worth utilities within each attribute and among the groups.

**Figure 1:**
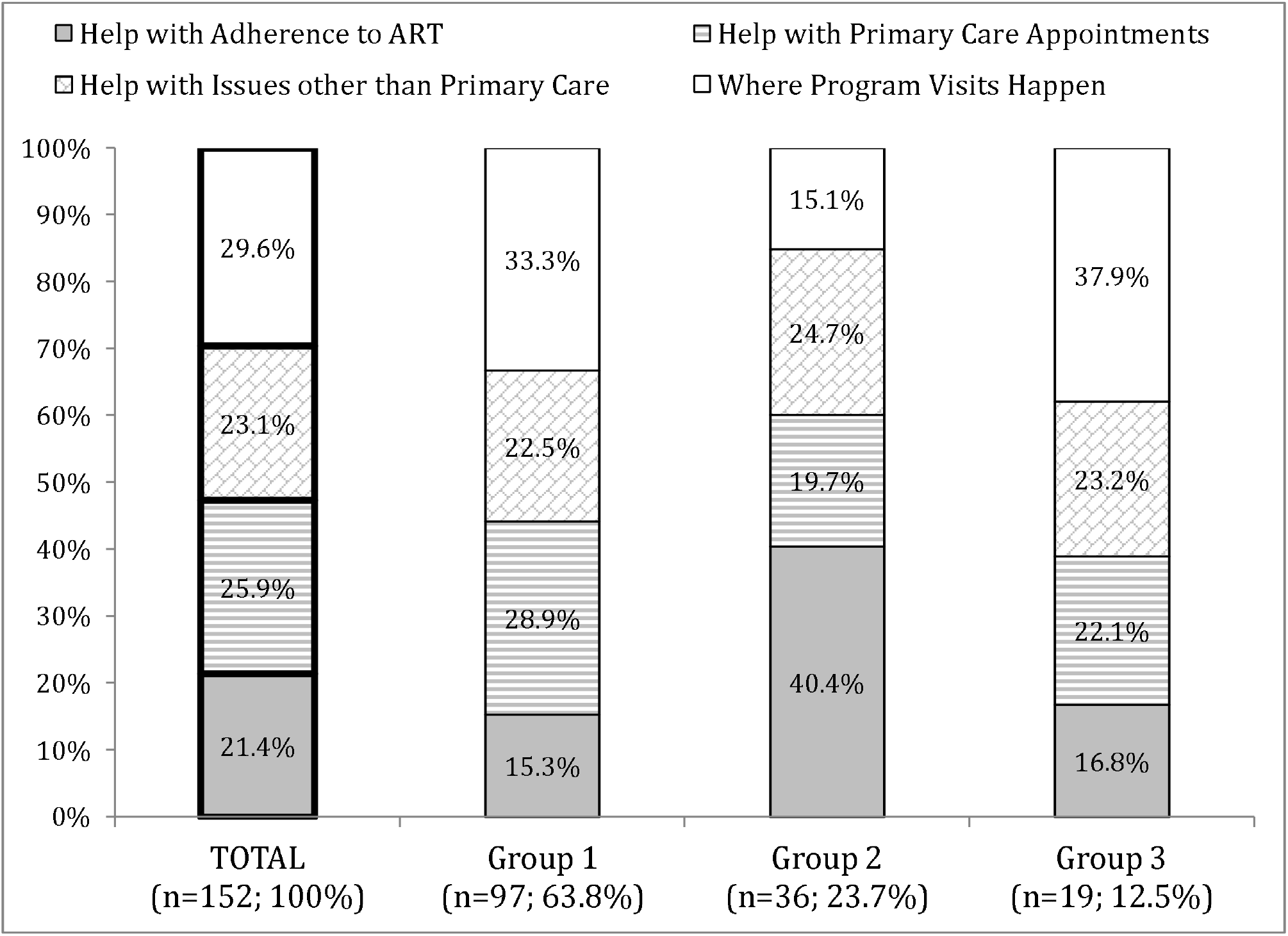
Relative importance of each attribute of the HIV Care Coordination Program for each provider group.

**Table 4:** Part-worth utilities (zero-centered values)

|  | Total<br>(n=152) | Group 1<br>(n=97) | Group 2<br>(n=36) | Group 3<br>(n=19) |
| --- | --- | --- | --- | --- |
| **Help with Adherence to ART** |  |  |  |  |
| Clients receive DOT or modified DOT | 31.09 | 28.99 | 75.07 | -41.51 |
| Clients receive medication reminders by phone call or text | -9.45 | -24.02 | 11.49 | 25.25 |
| Clients don't receive medication reminders, but are assessed and helped with medication adherence | -21.64 | -4.97 | -86.56 | 16.27 |
| **Help with Primary Care Appointments** |  |  |  |  |
| Staff provide reminders and attend all primary care appointments with clients | 29.71 | 47.35 | 21.42 | -44.60 |
| Staff provide reminders and arrange transportation for clients to get to primary care appointments | 19.93 | 20.49 | 28.63 | 0.64 |
| Staff only provide reminders for primary care appointments | -49.65 | -67.83 | -50.05 | 43.97 |
| **Help with Issues other than Primary Care** |  |  |  |  |
| Staff help with insurance, SSI benefits, and other general paperwork for health care coverage and benefits | 2.14 | -4.44 | 11.99 | 17.12 |
| Staff help with securing housing and food | -50.13 | -46.76 | -51.81 | -64.13 |
| Staff help with mental health and well-being issues (such as stress, substance use, diet, or personal relationships) | 20.75 | 9.64 | 47.16 | 27.44 |
| Staff help with connections to specialty medical care (cardiology, oncology, neurology, ear-nose-throat, etc.) | 27.23 | 41.57 | -7.35 | 19.56 |
| **Where Program Visits Happen** |  |  |  |  |
| Staff meet with clients at the program location | 3.39 | -7.03 | -5.27 | 72.98 |
| Staff meet with clients by phone or video chat | -44.30 | -63.75 | -24.09 | 16.70 |
| Staff make home visits, 30 minutes from the program location | 8.76 | 3.09 | 34.58 | -11.19 |
| Staff make home visits, 60 minutes from the program location | 32.15 | 67.69 | -5.23 | -78.49 |

**Figure 2:**
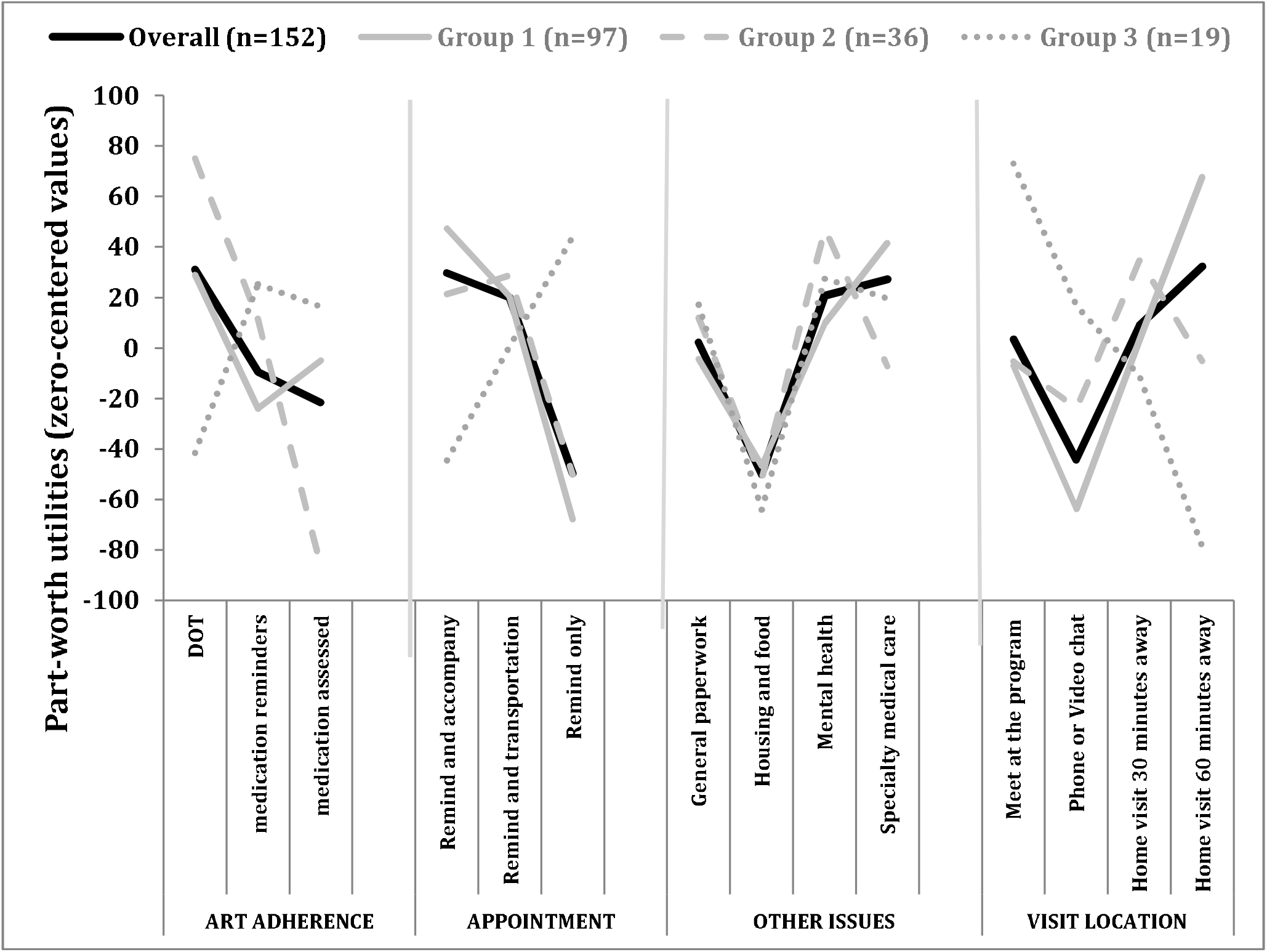
Plot of part-worth utilities for each level by provider group.

For the total sample of the providers, *Visit Location* had the highest relative importance (29.6%), followed by *Primary Care Appointments* (25.9%), *Issues other than Primary Care* (23.1%), and *Adherence to ART* (21.4%). Similarly, for the Group 1 providers, *Visit Location* was also the most important attribute (33%), followed by *Primary Care Appointments* (28.9%), *Issues other than Primary Care* (22.5%), and *Adherence to ART* (15.3%). In contrast, for the Group 2 providers, *Adherence to ART* was the most important attribute (40.4%), followed by *Issues other than Primary Care* (24.7%), *Primary Care Appointments* (19.7%), and *Visit Location* (15.1%). Finally, for the Group 3 providers, *Visit Location* was the most important attribute (37.9%), followed by *Issues other than Primary Care* (23.2%), *Primary Care Appointments* (22.1%), and *Adherence to ART* (16.8%). In summary, *Visit Location* had the highest importance for Group 1 (the largest group), Group 3, and overall, but had the lowest importance for Group 2. On the other hand, *Adherence to ART* had the highest importance for Group 2, but had the lowest importance for the other two groups and overall.

For the total sample, the levels within each attribute providers endorsed were *Clients receive DOT* for the *Adherence to ART* attribute, *Remind and Accompany* for the *Primary Care Appointment* attribute, *Special Medical Care* for the *Issues other than Primary Care* attribute, and *Home visit 60 minutes away* for the *Visit Location* attribute. Providers in Group 1 had the same pattern of preferences as for the sample as a whole. Group 2 providers also endorsed *Clients receive DOT*, but had the highest preferences for *Remind and Transportation, Mental Health Care*, and *Home Visit 30 minutes away*. Although the Group 1 and Group 3 providers have a similar pattern of importances, Group 3 providers’ part-worth utilities were very different from that of Group 1 providers. Providers from Group 3 endorsed *Medication reminders, Remind only, Mental health Care*, and *Meet at the program*.

Figure 2 displays the patterns of part-worth utilities within an attribute by groups. For example, the pattern of the part-worth utilities for *Issues other than Primary Care* Attribute was very similar across the 3 groups. In contrast, for the *Adherence to ART* attribute, the pattern for Group 2 providers deviated from the other two groups, and for the *Primary Care Appointments* and *Visit Location* attributes, the pattern for Group 3 providers deviated from the other two groups of providers.

### Preferences for Care Coordination Program Features

Using the choice simulation analysis, we estimated shares of preference for eight hypothetical programs in a single simulation scenario. We aggregated the shares of preference for programs of similar intensity from Table 2: Basic medical case management, Medium level medical case management, and Intensive medical case management. Figure 3 shows the shares of preference for the aggregated hypothetical CCPs for all providers and by LCA group. For the total sample of providers, the Intensive medical case management program which included *Mental health care* or *Specialty medical care* with *Home visits 30 or 60 minutes* from the program location had the highest degree of endorsement (62%). These programs also had the highest share for Group 2 (82%) and Group 1 (64%) providers. In contrast, Group 3 providers tended to endorse Basic medical management programs (69%).

**Figure 3:**
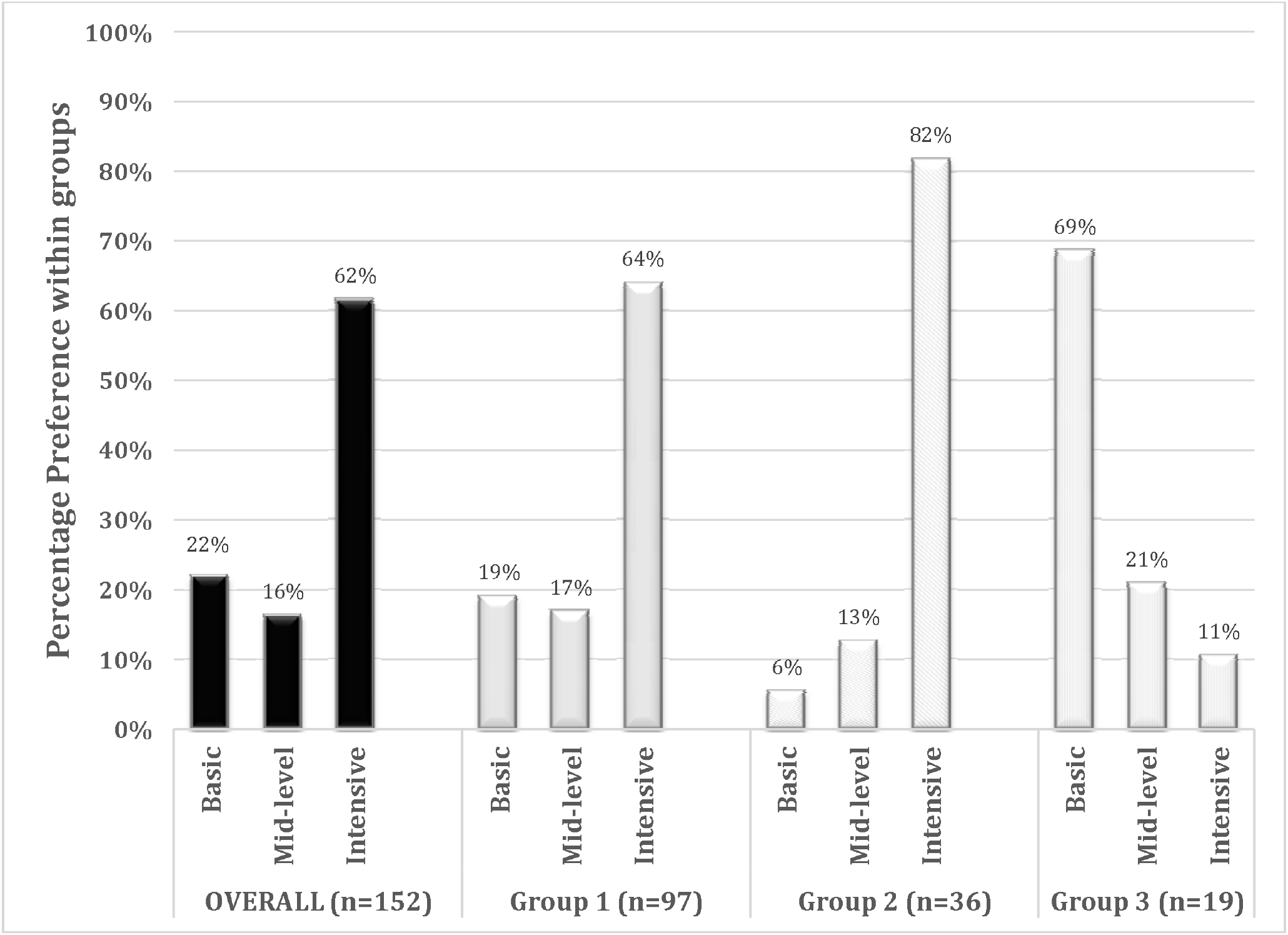
Choice simulation of the eight hypothetical care coordination programs, overall and by group.

## Discussion

In this study we sought to understand and to describe the heterogeneity of provider preferences for discrete features of an HIV care coordination program in NYC. Our findings indicate that the revised CCP program’s flexibility allows providers to deliver services both between and within agencies in a variety of ways, under a variety of conditions, to a variety of clients, and suggest some possible ways to improve fidelity to the core characteristics of the revised CCP.

Our analysis revealed underlying differences in preferences among providers, in particular within *Visit location* and *Adherence to ART*. Though only agency type was a statistically significant characteristic between groups, it may nonetheless be relevant that Group 3 had the highest proportion of providers who worked at an agency in Queens compared to Groups 1 and 2, and the highest proportion of providers who worked at an agency newly participating in CCP under the revised model. Queens has more “transit deserts” than other NYC boroughs, thus, geography may play a role in providers’ preferences for visit location [24]. Namely, providers may be more likely to endorse home visits if they work at agencies with access to multiple public transportation options or with sufficient home-visiting staff to cover a large geographic area between them [25,26]. In addition, staff at agencies with experience under the original and revised CCP may recognize the cumulative value of outreach work and home visits, despite the number of individual attempts that fail due to barriers such as clients not being home or not letting the provider in, compared to providers newly participating in the revised CCP.

While staff role was not a statistically significant characteristic between groups, a larger majority of staff in Group 3 were patient navigator-type staff (84.2%), compared to Groups 1 and 2 (64.9% and 55.6%, respectively). Staff in the various roles supporting CCPs may have different perspectives on DOT as an adherence strategy. In contrast to patient navigators, administrative staff do not themselves administer the DOT service, which requires substantial time and effort to deliver. Administrators often act as the liaisons between their programs and the NYC Health Department, and may be especially aware that the Health Department emphasizes DOT as an essential ART adherence strategy for the CCP. As a result, they may be more apt to endorse DOT as an effective and evidence-based programmatic approach to improve treatment adherence [27], without as much regard for the potential barriers to implementing DOT which may be more influential in patient navigator preferences.

The results from the choice simulation highlight the endorsement among most providers in the study for intensive CCPs that provide support for psychosocial factors that often act as barriers to ART adherence, such as mental healthcare and securing stable housing; this finding is substantially supported by past research [28–31]. A small subset of providers had a high share of preference in the simulation for Medical management-focused CCP rather than Intensive CCP. Though not statistically significant in our small study, heterogeneity is expected in a large city with diverse client needs and widely spread Ryan White Part A agencies. Therefore, this finding may indicate the need to facilitate additional opportunities to share lessons learned among agencies as well as across roles within agencies.

There are limitations to this study. First, though the sample size was within range for an unstratified conjoint study, we were unable to detect statistically significant differences between groups in our segmentation analysis [19]. In addition, data collected in DCEs are stated preferences, not actual provider behavior. Our analysis also did not take into account differences in client populations at each agency that could influence providers’ approach to service delivery. Unmeasured characteristics of the client populations may have influenced the ranges in preferences between groups. The interpretation of the findings is limited by the framing of the survey; it is unclear if providers’ stated preferences were based on what they prefer for themselves (to deliver) or for clients (to receive). Regardless of the motivation behind the preferences, the provider preferences identified in the DCE effectively highlight the factors that engage providers in service delivery, which addresses the overall aim of the study. Lastly, because the surveys were completed by March 2020, they do not account for possible changes in provider preferences as a result of the COVID-19 pandemic.

## Conclusion

The goal of this study was to characterize the heterogeneity of provider preferences for features of the HIV CCP using a latent class analysis. Our results highlight the endorsement among most providers of a CCP with intensive program options around ART and visit location, as well as ways to help mitigate psychosocial barriers to adherence such as unstable housing and mental health challenges. Our results also support the revised CCP’s flexible service models that enable providers to meet the needs of the clients these programs serve, and suggest that additional information sharing across and within agencies may be warranted to improve the fidelity with which the CCP is implemented. Further research is needed to understand components of the CCP endorsed by clients.

## Data Availability

All data produced in the present study are available upon reasonable request to the authors

## Authors’ contributions

DN and MI conceptualized the study. ABL conducted formative work. ABL, DN, MI, and RZ collaborated on the design of the data collection tool. CF and RZ performed statistical analyses. CF, MC, and RZ wrote the first draft of the paper. CF, MC, RZ, ABL, MR, JC, GG, ABL, MR, MI, and DN contributed to interpreting the data and to the writing and revising of the manuscript.

## Additional acknowledgements

We would like to acknowledge Sarah Kulkarni for her help with logistics and securing funding for the study; Sarah Kozlowski for her help with logistics and recruitment; Kate Taylor for her contribution to the conceptualization and drafting of the DCE; and Graham Harriman and the PROMISE qualitative research team [Rachel Schenkel, Thamara Tapia, Miguel Hernandez and Honoria Guarino], for their contributions to the larger project. We would also like to acknowledge the PROMISE Study Advisory Board members for their contributions to the study (in alphabetical order by last name): Mohammed Aldhuraibi for ACACIA Network, Lori Hurley for the STAR Program at SUNY Downstate Medical Center, Tiffany Jules for Services for the UnderServed, Inc., Genesis Luciano for AIDS Center of Queens County, Cyndi Morales for the Council on Adoptable Children, and Vanessa Pizarro for COMPASS.

## Supplemental Materials

**Supplemental Table 1.**
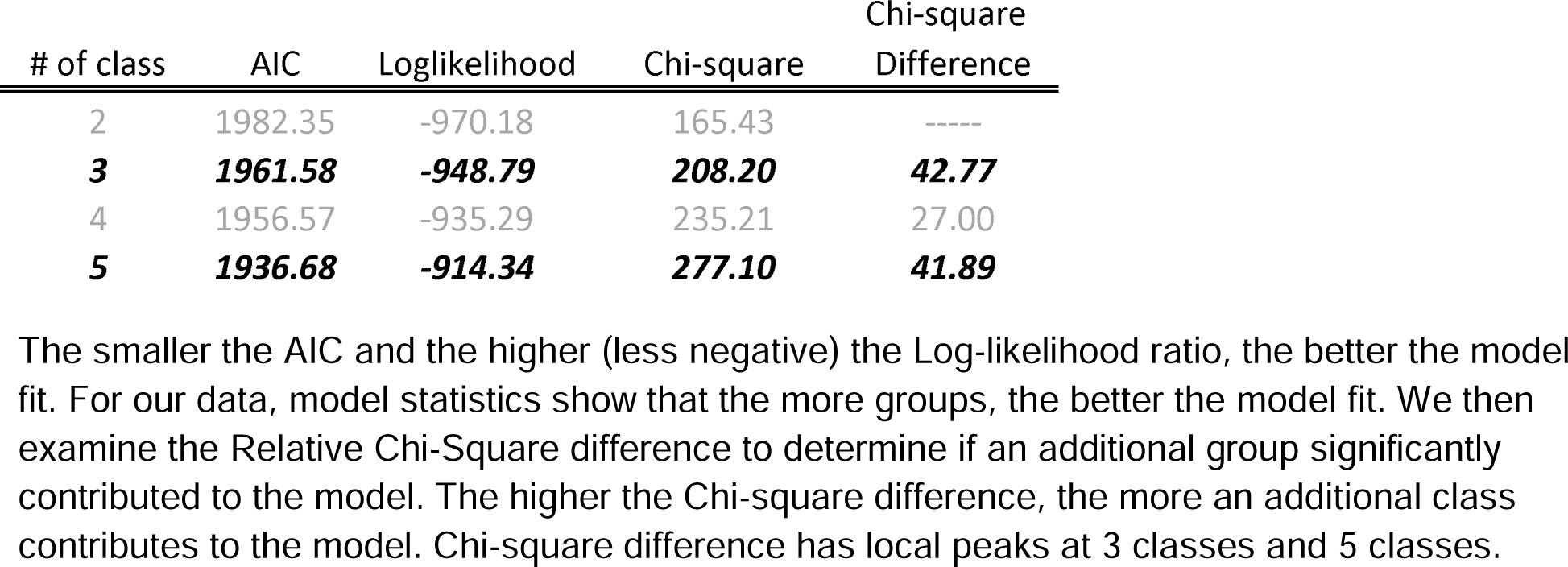
Comparing model estimates for different class solutions.

